# Abnormal liver tests in admitted patients with SARS-Cov-2 or other respiratory viruses- prognostic similarities and temporal disparities

**DOI:** 10.1101/2020.10.23.20218230

**Authors:** Noa Shafran, Assaf Issachar, Tzippy Shochat, Inbal Haya Shafran, Michael Bursztyn, Amir Shlomai

## Abstract

**Background and Aims:** Abnormal liver tests are common in patients with severe acute respiratory syndrome corona virus 2 (SARS-CoV-2) infection, but their association with short-term outcomes is controversial. We aimed to compare the pattern of abnormal liver tests in SARS-CoV-2 patients with those of patients infected with influenza or respiratory syncytial virus (RSV), two non-hepatotropic respiratory viruses, and their association with in-hospital mechanical ventilation or death.

**Methods:** A retrospective cohort study of 1271 hospitalized patients (872 influenza, 218 RSV, and 181 SARS-Cov-2) in a tertiary medical center. We defined abnormal liver tests as GPT, GOT or GGT≥40IU/ML at any time-point during hospitalization.

**Results:** Abnormal liver tests were mild-moderate in the majority of patients regardless of infection type but the majority of patients with influenza or RSV had a transaminases peak earlier during hospitalization compared to patients with SARS-Cov-2. Abnormal liver tests correlated with markers of severe disease across all types of infections, and were associated with mechanical ventilation or death, occurring mainly in patients with severe liver tests abnormalities (>200IU/L) (27.2%, 39.4% and 55.6% of patients with influenza, RSV or SARS-Cov-2). In multivariate analysis, controlling for age, gender, lymphopenia and CRP, liver tests abnormalities remained significantly associated with mechanical ventilation or death for influenza (OR= 3.047, 95% CI 1.518-6.117) and RSV (OR= 3.402, 95% CI 1.032-11.220) but not for SARS-Cov-2 (OR= 0.995, 95% CI 0.198-4.989). These results were confirmed upon propensity score matching.

**Conclusions:** Abnormal liver tests during hospitalization with different viral respiratory infections are common, may differ in their time-course and reflect disease severity. They are associated with worse outcomes, mainly in patients with severe liver test abnormalities, regardless of infection type.

## Background

The SARS-Cov-2 virus is the causative agent of the current Coronavirus disease 2019 (COVID-19) pandemic, first emerging in China(1), but now affecting most countries in the world(2). Although the respiratory system is the main target of the virus, SARS-Cov-2 often triggers a systemic immune-mediated response that might result in injury to various organs, such as the kidneys, heart or liver(3, 4). Recent reports indicate that hepatic injury is not uncommon among patients with COVID-19(5-7), and further suggest that abnormal liver tests are associated with increased mortality and should therefore be carefully monitored during hospitalization(8-10). However, the predictive value and the association of abnormal liver tests with worse outcomes in patients with COVID-19 is still controversial(11, 12).

Interestingly, SARS-coronavirus-associated hepatitis has been reported well before the present pandemic(13). Furthermore, the liver might also be collaterally damaged by infections with other respiratory viruses, such as influenza(14). Although few reports have suggested that certain strains of influenza A in children are possibly hepatotropic and might promote liver injury by directly targeting the hepatocytes (15), the widespread concept favors an immune-mediated mechanism(16), medication related hepatotoxicity, or hypotension that might result in hepatic ischemic injury.

We speculate that liver injury in COVID-19 is rather a marker of disease severity as in other viral respiratory infections (17). Accordingly, we aimed to investigate the prevalence and natural course of abnormal liver tests, as well as their association with the composite outcome of mechanical ventilation or death, in patients with COVID-19 as compared to patients with influenza or respiratory syncytial virus (RSV) infections.

## Methods

### Study design and patients

This retrospective study is based on prospectively collected data of patients older than 18 years admitted to our medical center from October 2018 to mid-July 2020, found to be positive for either influenza (A/B), RSV or SARS-Cov-2 by RT-PCR. Positive RT-PCR patients for more than one of the examined viruses, or lacking full information on liver enzymes were excluded. Local Institutional Review Board (#RMC-20-0142) approved the study that was performed according to Helsinki declaration.

### Definitions and outcomes

We defined abnormal liver tests, including aspartate transaminase (GOT), alanine transaminase (GPT), or gamma-glutamyl transferase (GGT), as levels ≥ 40 (U/L). These values are close to the accepted values defined in the literature (https://gi.testcatalog.org). The primary outcome of the study was defined as death or mechanical ventilation during hospitalization.

### Data collection

The following data were prospectively collected from the electronic medical records of patients included in the study: Basic characteristics, such as age, gender, body mass index (BMI), date of admission and outcomes (mechanical ventilation, length of hospitalization, discharge, transfer to ICU or death). Background liver disease was defined as documented cirrhosis, viral hepatitis or fatty liver (either previously recorded in the electronic files or observed by imaging during hospitalization), and major cardio-vascular diseases were defined as known ischemic heart disease, hypertension or diabetes. Clinical parameters recorded during hospitalization, including minimal blood oxygen saturation (BOS_min_), maximal fever, minimal systolic blood pressure (SBP_min_), as well as the presence of in-hospital pneumonia, treatment with anti-viral or anti-microbial agents were collected from the electronic medical files. Laboratory data of liver enzymes were extracted at 4 time points: on admission, last test before discharge, as well as maximum and minimum levels during hospitalization. In some cases, these time points overlapped. Maximum and minimum levels of the following laboratory parameters were collected for correlation analyses: white blood cell (WBC) count, C-reactive protein (CRP), lactate dehydrogenase (LDH), lymphocytes, ALKP and liver synthetic functions.

Factors related to elevated liver enzymes, such as serology tests for CMV, EBV, HIV, Herpes Simplex Virus (HSV), Varicella Zoster Virus (VZV), hepatitis A, B, C, and relevant imaging tests during hospitalization (abdominal sonography or abdominal computed tomography) were extracted from the patients’ electronic files.

### Statistical analysis

We used SAS version 9.4 and PRISM 8 softwares. Normality tests were conducted upon all variables. Due to the non-normal distribution of our variables we used the non-parametric Wilcoxon and Kruskal Wallis tests when appropriate and median and interquartile range (IQR) are presented. Chi-square or Fisher’s exact tests were compared categorical variables between study groups, accordingly. Two-sided p<0.05 was considered statistically significant.

The patients cohort was divided into three RT-PCR based viral diagnosis groups and then further divided according to their liver enzyme status. Baseline characteristics, laboratory tests and basic information regarding hospitalization were compared between the groups and sub-groups. Univariate and multivariate regression models were constructed to estimate predictors for the primary outcome. Variables previously described found to be associated with the aforementioned outcomes, such as low lymphocyte counts, high CRP levels, age or gender (1, 18-20), as well as variables that were found to significantly associate in the univariate analyses, were incorporated into a Cox regression model. Odd ratios (ORs) with 95% confidence intervals (CI) were calculated. Logistic regression was used to calculate ORs.

As a sensitivity analysis, propensity score matching was used to identify a cohort of patient with similar risk factors and baseline characteristics that were shown to impact the outcome/used in the multivariate model. matching was performed with the use of a 1:1 matching protocol without replacement, and a regression model was performed on propensity-matched cohort of patients with and without abnormal liver tests for each virus independently. Analyses of the primary outcome was performed in a binary regression model.

## Results

### Patients’ characteristics according to infection type

We allocated all patients hospitalized in our medical center between October 2018 and mid-July 2020 due to infection with either influenza (A/B), RSV or SARS-Cov-2 (the later, during 2020 only). Overall, following exclusion of patients for various reasons, 1271 patients (872 influenza, 218 RSV, and 181 SARS-Cov-2) were included in the final analysis (Fig S1). Table 1 outlines the various demographics, as well as major clinical features and laboratory values. RSV patients were older with a significantly higher rate of cardiovascular diseases compared to those with either influenza or SARS-Cov-2 infections. In addition, a higher proportion of patients with RSV had pneumonia and accordingly, more of them were treated with antibiotics compared to the other two groups. The rates of mechanical ventilation or death, however, did not differ between the three infections.

**Table 1.** Major characteristics of patients according to the type of infection.

|  | Influenza<br>(N=872) | RSV<br>(N=218) | SARS-Cov-2<br>(N=181) | p Value |
| --- | --- | --- | --- | --- |
| Age (median, IQR) | 70.27 (51.82-81.01) | 75.26 (65.74-85.31) | 61 (45-72) | **<.0001** |
| Gender-females (%) | 55.96 | 51.83 | 43.6 | **0.009** |
| BMI (median, IQR) | 26.25 (22.67- 30.07) | 27.27 (23.58- 30.16) | 27.1 (24.5-31.2) | **0.012** |
| Prior liver disease (%) | 4.47 | 7.80 | 3.3 | 0.072 |
| Cardiovascular diseases | 38.65 | 55.05 | 43.1 | **<.001** |
| In-hospital pneumonia (%) | 17.43 | 22.94 | 3.3 | **<.001** |
| Antibiotic treatment (%) | 66.51 | 76.15 | 63.5 | **0.010** |
| Anti-viral treatment (%) | 82.00 | 39.45 | 33.7 | **<.001** |
| Saturation (%)<sub>min</sub> (median, IQR) | 95.00 (90.00-97.00) | 93.00( 90.00-95.00) | 94.00 (89.00- 96.00) | **<.001** |
| Systolic BP <sub>min</sub> (median, IQR) | 108.00 (95.00- 127.00) | 110.00 (96.50-125.50) | 110 (99-122) | 0.910 |
| Albumin <sub>min</sub> (median, IQR) | 3.60(3.20-4.00) | 3.50 ( 2.90- 4.00) | 3.1 (3.6-4.1) | 0.125 |
| Lymph-<sub>min</sub> (median, IQR) | 0.70 (0.40-1.00) | 0.70 (0.40-1.1) | 0.8 (0/6-1.25) | **<.0001** |
| CRP <sub>max</sub> (median, IQR) | 6.27 (2.96- 13.98) | 6.99 (3.63-15.60) | 6.9 (1.8-15.5) | 0.331 |
| GPT <sub>max</sub> (median, IQR) | 23.00 (16.00-37.50) | 22.50 (15.00-40.00) | 24.7 (15.5-45.2) | 0.383 |
| GOT<sub>max</sub> (median, IQR) | 30.00 (22.00-46.00) | 29.00 (21.00-48.00) | 29.8 (21.8-54.2) | 0.608 |
| ALKP<sub>max</sub> (median, IQR) | 82.00 (65.00-114.00) | 92.50 (67.00-129.00) | 81 (64-113.5) | 0.086 |
| GGT<sub>max</sub> (median, IQR) | 32.50 (18.00-75.50) | 43.00 (25.00-87.00) | 38.00 (21-81.5) | **0.001** |
| GPT <sub>max</sub> 40-200 (%) | 21.3 | 21.5 | 27.6 |  |
| GPT <sub>max</sub> 200-400 (%) | 1.1 | 2.29 | 2.2 |  |
| GPT <sub>max</sub> >400 (%) | 0.8 | 2.29 | 0.0 |  |
| GOT <sub>max</sub> 40-200 (%) | 28.4 | 25.2 | 30.4 |  |
| GOT <sub>max</sub> 200-400 (%) | 2.17 | 2.29 | 1.1 |  |
| GOT<sub>max</sub> >400 (%) | 1.03 | 2.7 | 0 |  |
| GGT<sub>max</sub> 40-200 (%) | 31.9 | 39.4 | 36.5 |  |
| GGT<sub>max</sub> 200-400 (%) | 5.5 | 7.7 | 7.7 |  |
| GGT<sub>max</sub> >400 (%) | 3.4 | 5 | 3.9 |  |
| TBil<sub>max</sub> (median, IQR) | 0.45 (0.30-0.70) | 0.55 (0.37-0.93) | 0.50 (0.34-0.73) | **<.0001** |
| INR<sub>max</sub> (median, IQR) | 1.09 (0.97-1.26) | 1.14 (1.01-1.48) | 1.1 (1.1-1.24) | **0.013** |
| Death (%) | 5.96 | 9.63 | 6.6 | 0.152 |
| Mechanical ventilation (%) | 3.67 | 5.96 | 6.6 | 0.110 |

GPT level was elevated in 23.2%, 26.1% and 29.8%, GOT was elevated in 31.6%, 30.2% and 31.5%, GGT was elevated in 40.8%, 52.1% and 48.1% of patients with influenza, RSV and SARS-Cov-2, respectively. Only ∼1-5% of patients had severe elevation in GPT or GOT (>200 IU/L) and ∼8-13% had severe elevation in GGT across all types of infections. Median values of GPT, GOT and ALKP did not significantly differ between the groups.

### The temporal pattern of abnormal liver tests according to the type of infection

In order to validate that the abnormal liver tests were by large transient and likely related to the acute illness, we compared the highest with the lowest values during hospitalization. There was a clear, and mostly statistically significant difference between the highest and the lowest levels of liver tests during hospitalization, for patients with abnormal values, across all types of viral infections (Fig S2).

We next analyzed the timing of peak abnormal liver enzymes during hospitalization (Fig 1). The GPT peak was recorded by day 6 in > 60% of patients with influenza or RSV infections, whereas only ∼40% of patients with SARS-Cov-2 peaked by day 6. Regarding GOT peak, >70% of patients with influenza or RSV infection reached their peak by day 6, whereas only ∼50% of patients with SARS-Cov-2 did.

**Fig 1.**
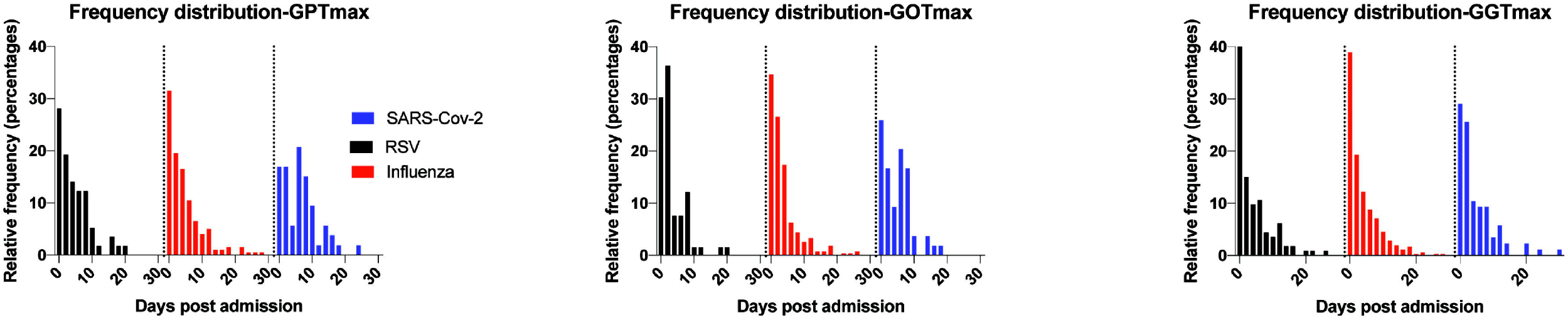
Time course of elevated liver tests in RSV, influenza or SARS-Cov-2 infections. Patients with eLFTs during hospitalization were allocated and the timing of their maximal value of LFT (days post admission) was plotted on a frequency distribution plot for each type of infection (influenza, RSV or SARS-Cov-2).

### Characteristics of patients and disease severity as a function of liver tests

Patients’ demographics and the severity of their disease according to liver tests were subsequently analyzed (Table 2). Influenza or RSV infected patients with abnormal liver tests had a higher rate of prior liver disease (6.7% vs. 2.1%, p=0.002 for influenza and 11.4% vs. 3.2%, p=0.039 for RSV). Such differences were not detected among patients with SARS-Cov-2. No significant difference in BMI between patients with or without abnormal liver tests was detected across all types of infections.

**Table 2.** Major characteristics and outcomes of patients according to the presence or absence of elevated LFTs (GPTmax ≥40, GOTmax ≥40 or GGTmax ≥40).

| Parameter | Influenza |  |  | RSV |  |  | SARS-Cov-2 |  |  |
| --- | --- | --- | --- | --- | --- | --- | --- | --- | --- |
|  | Elevated LFTs<br>N=450 | Normal LFTs<br>N=422 | p Value | Elevated LFTs<br>N=123 | Normal LFTs<br>N=95 | p Value | Elevated LFTs<br>N=103 | Normal LFTs<br>N=78 | p Value |
| Age | 71.86 (59.42-81.84) | 67.16 (36.18-80.71) | **0.0001** | 72.12 (63.75-82.91) | 79.91 (66.87-85.85) | **0.0242** | 61 (46-73) | 60.5 (42-70) | 0.654 |
| Gender female % | 48.00 | 64.45 | **<.001** | 51.22 | 52.63 | 0.892 | 31.1 | 60.3 | **<.001** |
| BMI (median, IQR) | 26.26 (22.66-30.04) | 25.86 (22.86-30.08) | 0.7783 | 26.86 (24.03-30.10) | 27.73 (22.49-30.39) | 0.5512 | 27.78 (24.8-32.4) | 26.3 (24-29.57) | 0.051 |
| Prior liver disease (%) | 6.67 | 2.13 | **0.002** | 11.38 | 3.16 | **0.039** | 2.9 | 3.8 | 0.728 |
| Major illness (DM, HTN)(%) | 46.00 | 30.81 | **<.001** | 52.85 | 57.89 | 0.494 | 41.7 | 44.9 | 0.674 |
| In-hospital pneumonia (%) | 21.33 | 13.27 | **0.0015** | 26.02 | 18.95 | 0.2297 | 5.8 | 0 | **0.017** |
| Antibiotics treatment (%) | 76.00 | 56.40 | **<.001** | 78.86 | 72.63 | 0.337 | 76.7 | 46.2 | **<.0001** |
| Anti-viral treatment (%) | 83.11 | 80.81 | **<.001** | 40.65 | 37.89 | 0.780 | 42.7 | 21.8 | **0.003** |
| Systolic BP (min) (median, IQR) | 110.00 (94.00-125.00) | 108.00 (95.00-129.00) | 0.4158 | 103.00 (90.00-120.00) | 117.00 (105.00-132.00) | **<.0001** | 106 (98-120) | 113 (100-127) | **0.045** |
| Saturation% (min) (median, IQR) | 93.00 (89.00-96.00) | 96.00 (92.00-97.00) | **<.0001** | 92.00 (88.50-94.00) | 94.00 (92.00-96.00) | **0.0023** | 92 (87-95) | 95.00 (92.00-97.00) | **<.0001** |
| Albumin (mg/dL) (median, IQR) | 3.40 (2.80-3.80) | 3.70 (3.40-4.10) | **<.0001** | 3.25 (2.60-3.70) | 3.70 (3.30-4.10) | **<.0001** | 3.40 (2.80-3.80) | 3.9 (3.375-4.4) | **<.0001** |
| Lymp. min (median, IQR) | 0.60 (0.40- | 0.70 (0.50- | **<.0001** | 0.50 (0.30- | 0.90 (0.60- | **<.0001** | 0.70 (0.50- | 1.10 (0.80- | **<.0001** |
|  | 0.90) | 1.00) |  | 0.80) | 1.20) |  | 1.00) | 1.50) |  |
| CRP<sub>max</sub> (median, IQR) | 9.28 (4.06-19.45) | 4.64 (2.46-9.03) | **<.0001** | 10.32(4.25-18.96) | 4.86 (2.75-10.88) | **<.0001** | 11.95 (4.58-21.3) | 2.6 (1.07-7.07) | **<.0001** |
| GPT<sub>max</sub> (median, IQR) | 36.00 (24.00-57.00) | 16.50 (13.00-21.00) | **<.0001** | 36.00 (21.00-67.00) | 15.00 (13.00-20.00) | **<.0001** | 41.8 (26-78.7) | 16.9 (12.375-21.375) | **<.0001** |
| GOT<sub>max</sub> (median, IQR) | 45.00 (31.00-73.00) | 23.00 (18.00-29.00) | **<.0001** | 41.00 (27.00-75.00) | 22.00 (18.00-26.00) | **<.0001** | 42.35 (30-78.875) | 23 (19.60-26.90) | **<.0001** |
| ALKP<sub>max</sub> (median, IQR) | 95.50 (73.00-140.00) | 74.00 (60.00-93.00) | **<.0001** | 111.00 (82.00-155.00) | 74.00 (56.00-95.00) | **<.0001** | 87 (69-149) | 74.5 (59-92.25) | **<.0001** |
| GGT<sub>max</sub> (median, IQR) | 73.00 (43.00-152.00) | 19.00 (13.00-26.00) | **<.0001** | 77.00 (51.00-159.00) | 23.00 (16.00-31.00) | **<.0001** | 77.00 (48-185) | 19 (14-28) | **<.0001** |
| TBil<sub>max</sub> (mg/dL) (median, IQR) | 0.52 (0.34-0.83) | 0.39 (0.26-0.56) | **<.0001** | 0.65 (0.43-1.07) | 0.49 (0.32-0.80) | **0.0004** | 0.51 (0.40-0.86) | 0.41 (0.30-0.66) | **0.005** |
| INR<sub>max</sub> (median, IQR) | 1.16 (1.01-1.36) | 1.04 (0.95-1.14) | **<.0001** | 1.19 (1.04-1.56) | 1.05 (0.99-1.39) | **0.0396** | 1.2 (1.1-1.4) | 1.05 (1-1.105) | **<.0001** |
| Death (%) | 10.22 | 1.42 | **<.001** | 13.01 | 5.26 | 0.065 | 9.7 | 2.6 | 0.056 |
| Mechanical ventilation (%) | 6.00 | 1.18 | **<.001** | 10.57 | 0 | **<.001** | 10.7 | 1.3 | **0.012** |
| ICU admission (%) | 4.00 | 0.71 | **0.001** | 6.50 | 0 | **0.010** | 8.7 | 0 | **0.007** |
| Length of hospital stay (days) | 4.50 (3.00-9.00) | 3.00 (2.00-5.00) | **<.0001** | 7.00 (3.00-14.00) | 3.00 (2.00-6.00) | **<.0001** | 8 (4-12.5) | 5 (2-8) | **0.001** |

Patients with either influenza or SARS-Cov-2 with liver test abnormalities had a higher rate of pneumonia (21.3% vs. 13.3%, p=0.0015, 5.8% vs. 0.0%, p=0.017, respectively), a use of antibiotics (76% vs. 56.4%, p<0.001, 76.7% vs. 46.2%, p<0.0001, respectively) and anti-viral agents (83.11% vs. 80.81%, p<0.001, 42.7% vs. 21.8%, p=0.003, respectively) during hospitalization, compared to patients with normal enzymes.

There were significant differences in clinical and laboratory parameters of disease severity, including BOS_min_, CRP_max_, lymphocytes _min_, Albumin _min_ between patients with or without liver test abnormalities, regardless of infection type.

Consistently, patients with liver test abnormalities had a higher rate of mechanical ventilation (6.0% Vs 1.2%, p<0.001 for influenza, 10.6% vs. 0.0%, p<0.001 for RSV and 10.7% vs. 1.3%, p=0.012 for SARS-Cov-2), admission to ICU (4.0% vs. 0.7%, p=0.001 for influenza, 6.5% vs. 0%, p=0.010 for RSV and 8.7% vs. 0.0%, p=0.007 for SARS-Cov-2), and death (10.2% vs. 1.4%, p<0.001 for influenza, 13.0% vs. 5.3%, p=0.065 for RSV and 9.7% vs. 2.6%, p=0.056 for SARS-Cov-2), as well as a longer length of hospitalization days (4.5 (3-9) vs. 3 (2-5), p<0.001 for influenza, 7 (3-14) vs. 3 (2-6), p<0.001 for RSV and 8.0 (4-12.5) vs. 5.0 (2-8), p=0.001 for SARS-Cov-2), compared to patients with normal enzymes.

### Association of patients’ characteristics and parameters of disease severity with poor outcomes

A univariate analysis to test possible association of patients’ demographic, clinical or laboratory parameters with a composite outcome of mechanical ventilation or death during hospitalization was performed., Patients’ age, use of antibiotics, hospitalization length and admission to ICU as well as other disease severity parameters, including BOS_min_ and SBP_min_, were associated with the composite outcome in all types of infection (Table S1).

Routinely used laboratory parameters to evaluate disease severity, CRP and albumin levels, were associated with the composite outcome in all three types of infection. Importantly, the same was true for liver tests (GPT, GOT or GGT), all strongly associated with the composite outcome in the three groups.

Incorporated liver test abnormalities in a multivariate regression analysis utilizing patients’ age and gender as well as CRP and lymphocyte count (Fig 2), were found to be significantly associated with the composite outcome in patients with either influenza or RSV infections (OR= 3, 95% CI 1.518-6.117 for influenza and OR= 3.4, 95% CI 1.032-11.220 for RSV), but not in patients with SARS-Cov-2 (OR= 0.995, 95% CI 0.198-4.989).

**Fig 2.**
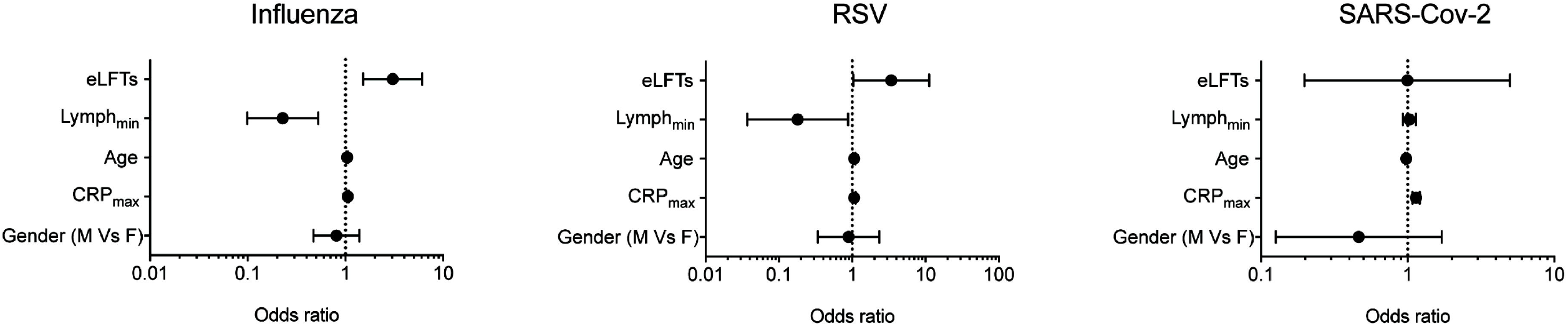
A multivariate analysis of various parameters related to patients’ basic demographics and disease severity and their association with a composite outcome of death or mechanical ventilation. The odds ratios (OR) for each parameter incorporated in the analysis and 95% confidence limits are plotted (eLFTs-elevated liver function tests, M-males, F-females).

We also performed the regression model on propensity-matched cohort of patients with similar baseline characteristics that were shown to impact the outcome, with abnormal liver tests versus normal liver tests. Primary outcome, defined as death or intubation was associated with abnormal liver tests in influenza and RSV (OR= 5.3, 95% CI 2.398-11.726, p<0.001 for influenza and OR= 5.8, 95% CI 1.618-21.03 p=0.007 for RSV), but not with SARS-Cov-2 (OR= 2.4, 95% CI 0.512-11.511 p=0.264).

We next plotted the peak levels of GOT, GPT or GGT recorded during hospitalization against minimal albumin or maximal CRP values for each patient in our cohort (Figs S3,S4). There was an inverse statistically significant correlation between liver tests and albumin levels and a linear statistically significant correlation between liver tests and CRP levels for GOT, GPT or GGT. This suggests that liver test abnormalities correlate well with other markers of disease severity.

### Correlation of patients’ poor outcomes with the severity of liver test abnormalities

Percentage of patients who achieved the composite outcome correlated well with the severity of liver tests abnormalities (Fig 3A). We categorized patients as having normal liver enzymes (GPT, GOT and GGT<40IU/L), mild to moderate liver test abnormalities (at least one enzyme ≥40IU/L but <200IU/L) or severe liver test abnormalities (at least one of the enzymes ≥200IU/L). The percentage of patients with the composite outcome was 2.6%, 9.6% and 27.2% (p<0.001) for influenza, 4.3%, 13.0% and 39.4% (p<0.001) for RSV and 0.0%, 3.7% and 55.6% (p<0.001) for SARS-Cov-2, respectively. Consistently, GPT_max_ GOT_max_ and GGT_max_ levels of patients with the composite outcome were significantly higher compared to all other patients, regardless of infection type (Fig 3B).

**Fig 3.**
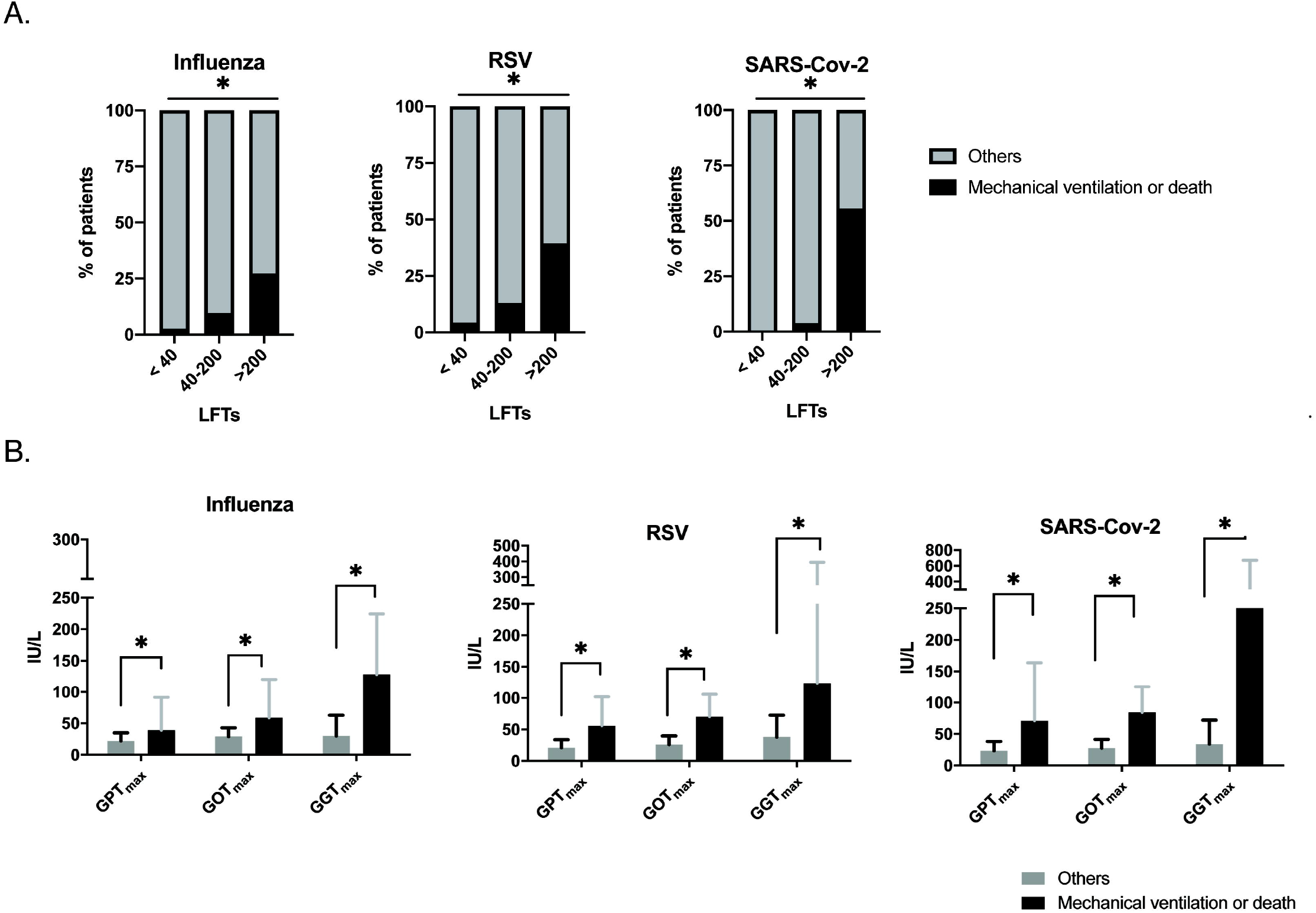
Association of disease severity with the magnitude of liver tests abnormalities. **A**. Stacked graphs presenting the percentage of patients that either achieved or not achieved (others) the composite endpoint (mechanical ventilation or death) as a function of their LFTs (GPT, GOT and GGT <40IU/L, at least one of the enzymes> 40IU/L but <200IU/L or at least one of the enzymes >200IU/L) for influenza, RSV or SARS-Cov-2 infections (*p<0.001). **B**. Median values and range (upper/lower limits) of LFTs for patients that either achieved or not (others) the composite endpoint (mechanical ventilation or death) for influenza, RSV or SARS-Cov-2 infections (*p<0.001).

## Discussion

Here, we show that abnormal liver tests are common among admitted patients with either influenza, RSV or SARS-Cov-2 infections, and that the percentage of patients with abnormal liver tests at some point during infection does not differ much between infections, ranging from ∼25-30% (for transaminases elevation) to nearly 50% (for GGT elevation) of patients. Several recent studies have highlighted the relatively high frequency of abnormal liver tests among patients infected with SARS-Cov-2(6, 21). Our study suggests that this phenomenon is not unique to this infection but rather similarly occurs within other major respiratory viral infections. However, the kinetics of abnormal liver tests may differ between infections, with a delayed elevation of liver tests in the case of SARS-Cov-2 compared to influenza or RSV, compatible with the well-established observation that clinical deterioration of SARS-Cov-2 patients is typically delayed until days 7-10 after symptom onset (22).

Disease severity or worse outcomes parameters were significantly more common among patients with abnormal liver tests regardless of the type of infection, in line with other parameters, both clinical or laboratory, known to be associated with disease severity.

Therefore, it is reasonable to speculate, that abnormal liver tests are a surrogate marker of disease severity and worse outcomes in patients hospitalized due to acute respiratory viral infection, and that this is not a property unique to a particular virus. In support of this, liver enzymes were all found to be inversely correlated with the minimal value of serum albumin and linearly correlated with maximal CRP levels recorded during hospitalization, two other well-established markers for disease severity.

Our study indicates that patients with the highest risk of composite outcome are those with severe abnormal liver tests (>200 IU/L) in all types of infections, in line with a recent report from a large cohort of SARS-Cov-2 patients in the US, indicating that in the 6.4% of patients with severe abnormal liver tests (>5 times normal range), worse outcomes should be anticipated (23). Still, the majority of our patients with abnormal liver tests presented with only mild-moderate liver enzymes abnormalities.

Several mechanisms may explain liver biochemical abnormalities during non-hepatotropic viral infection, but the leading theory is that liver injury is immune mediate, resulting from molecular mimicry between viral and hepatocyte antigens(16). However, a recent study has suggested that SARS-Cov-2 contributes to hepatic impairment by directly infecting the liver resulting in a typical histological signature (24). These authors related relatively mild alterations in transaminase levels to hepatic impairment, reflected by a low albumin level, and to massive apoptosis observed in few liver biopsies. Nevertheless, this study has recently been challenged (25, 26), arguing that the relatively mild abnormal liver tests do not reflect true liver injury, usually defined by a much higher elevations in transaminases (27). Moreover the observed low albumin levels could reflect the severity of the disease rather than liver synthetic dysfunction. Indeed, our study suggests that abnormal liver tests correlate well with low albumin level as well as with other markers of disease severity, such as CRP and that in most infected patients with abnormal liver tests, the impairment is mild and is not virus specific.

Although this study includes a relatively large number of patients with an ample of clinical and laboratory data, it also has several limitations: First, it is retrospective and not all patients were systematically tested for liver enzymes and we had no control on the timing by which these tests were taken for each patient. Second, we had no access to laboratory results prior to hospitalization and could not accurately assess whether the observed abnormal liver tests during hospitalization is entirely new. We partially overcome this pitfall, by showing that the peak in abnormal liver tests is significantly higher than the nadir during hospitalization, suggesting that at least in the majority of patients, the maximal abnormal liver tests values are related to the acute disease and not to an underlying liver problem. Third, the cutoff value of 40IU/L for a definition of elevated GPT, GOT and GGT levels is largely arbitrary and was not adjusted for age, weight and gender. However, these cutoffs were used in previous studies(24), and by and large represent values in the range observed in 95% of the healthy population. In addition, the reason that we have not included the ALKP values in our primary analysis is that this enzyme is not necessarily liver-specific.

In summary, in this study we show that abnormal liver tests are quite common and appear with similar frequency among hospitalized patients with either influenza, RSV or SARS-Cov-2 infections, although the timing of abnormal liver tests peak may be somewhat delayed in patients with SARS-Cov-2. The abnormal liver tests correlate with other markers of disease severity in all three types of infection and are associated with worse outcomes. Although our results suggest that abnormal liver tests do not necessarily reflect a direct and significant liver injury, but is rather a consequence of a systemic inflammatory response, we believe that liver tests should be routinely taken in such hospitalized patients, and that abnormal values should alert the treating physician to monitor the patient more intensively.

## Data Availability

The data will be available upon request

## List of abbreviations

(SARS-CoV-2): Severe acute respiratory syndrome corona virus 2
(RSV): respiratory syncytial virus
(COVID-19): Coronavirus disease 2019
(GOT): aspartate transaminase
(GPT): alanine transaminase
(GGT): gamma-glutamyl transferase

## Notes

**Conflict of interest statement:** The authors declare no conflict of interest regarding this Work

### Competing Interest Statement

The authors have declared no competing interest.

### Funding Statement

There is no funding relevant to this publication

### Author Declarations

IRB, Rabin Medical Center

## References

1. Guan W-j, Ni Z-y, Hu Y, Liang W-h, Ou C-q, He J-x, et al. Clinical Characteristics of Coronavirus Disease 2019 in China. New England Journal of Medicine 2020.

2. Roser M, Ritchie H, Ortiz-Ospina E. Coronavirus Disease (COVID-19) In. Published online at OurWorldInData.org.; 2020.

3. Chen N, Zhou M, Dong X, Qu J, Gong F, Han Y, et al. Epidemiological and clinical characteristics of 99 cases of 2019 novel coronavirus pneumonia in Wuhan, China: a descriptive study. Lancet 2020;395:507–513.

4. Huang C, Wang Y, Li X, Ren L, Zhao J, Hu Y, et al. Clinical features of patients infected with 2019 novel coronavirus in Wuhan, China. Lancet 2020;395:497–506.

5. Mao R, Qiu Y, He J-S, Tan J-Y, Li X-H, Liang J, et al. Manifestations and prognosis of gastrointestinal and liver involvement in patients with COVID-19: a systematic review and meta-analysis. The Lancet Gastroenterology & Hepatology 2020:1–13.

6. Schaefer EAK, Arvind A, Bloom PP, Chung RT. Interrelationship Between Coronavirus Infection and Liver Disease. Clinical Liver Disease 2020;15:175–180.

7. Xie H, Zhao J, Lian N, Lin S, Xie Q, Zhuo H. Clinical characteristics of non-ICU hospitalized patients with coronavirus disease 2019 and liver injury: A retrospective study. Liver International 2020;40:1321–1326.

8. Hundt MA, Deng Y, Ciarleglio MM, Nathanson MH, Lim JK. Abnormal Liver Tests in COVID-19: A Retrospective Observational Cohort Study of 1827 Patients in a Major U.S. Hospital Network. Hepatology 2020; doi: 10.1002/HEP.31487.

9. Lei F, Liu YM, Zhou F, Qin JJ, Zhang P, Zhu L, et al. Longitudinal association between markers of liver injury and mortality in COVID-19 in China. Hepatology (Baltimore, Md.) 2020:hep.31301-31323.

10. Qi X, Liu C, Jiang Z, Gu Y, Zhang G, Shao C, et al. Multicenter analysis of clinical characteristics and outcome of COVID-19 patients with liver injury. Journal of hepatology 2020:1–14.

11. Romana Ponziani F, Del Zompo F, Nesci A, Santopaolo F, Ianiro G, Pompili M, et al. Liver involvement is not associated with mortality: results from a large cohort of SARS-CoV-2 positive patients. Aliment Pharmacol Ther 2020.

12. Vespa E, Pugliese N, Piovani D, Capogreco A, Danese S, Aghemo A. Liver tests abnormalities in COVID-19: trick or treat? J Hepatol 2020.

13. Chau TN, Lee KC, Yao H, Tsang TY, Chow TC, Yeung YC, et al. SARS-associated viral hepatitis caused by a novel coronavirus: report of three cases. Hepatology 2004;39:302–310.

14. Papic N, Pangercic A, Vargovic M, Barsic B, Vince A, Kuzman I. Liver involvement during influenza infection: perspective on the 2009 influenza pandemic. Influenza and Other Respiratory Viruses 2011;6:e2–e5.

15. Whitworth JR, Mack CL, O’Connor JA, Narkewicz MR, Mengshol S, Sokol RJ. Acute hepatitis and liver failure associated with influenza A infection in children. J Pediatr Gastroenterol Nutr 2006;43:536–538.

16. Adams DH, Hubscher SG. Systemic viral infections and collateral damage in the liver. Am J Pathol 2006;168:1057–1059.

17. Thorburn K, Fulton C, King C, Ramaneswaran D, Alammar A, McNamara PS. Transaminase levels reflect disease severity in children ventilated for respiratory syncytial virus (RSV) bronchiolitis. Scientific Reports 2018:1–6.

18. Bhargava A, Fukushima EA, Levine M, Zhao W, Tanveer F, Szpunar SM, et al. Predictors for Severe COVID-19 Infection. Clinical Infectious Diseases 2020.

19. Chen T, Wu D, Chen H, Yan W, Yang D, Chen G, et al. Clinical characteristics of 113 deceased patients with coronavirus disease 2019: retrospective study. Bmj 2020;368:m1091.

20. Zhou F, Yu T, Du R, Fan G, Liu Y, Liu Z, et al. Clinical course and risk factors for mortality of adult inpatients with COVID-19 in Wuhan, China: a retrospective cohort study. Lancet 2020;395:1054–1062.

21. Bertolini A, van de Peppel IP, Bodewes FAJA, Moshage H, Fantin A, Farinati F, et al. Abnormal liver function tests in COVID-19 patients: relevance and potential pathogenesis. Hepatology 2020; doi: 10.1002/hep.31480

22. Zhou F, Yu T, Du R, Fan G, Liu Y, Liu Z, et al. Clinical course and risk factors for mortality of adult inpatients with COVID-19 in Wuhan, China: a retrospective cohort study. The Lancet 2020;395:1054–1062.

23. Phipps MM, Barraza LH, LaSota ED, Sobieszczyk ME, Pereira MR, Zheng EX, et al. Acute Liver Injury in COVID-19: Prevalence and Association with Clinical Outcomes in a Large US Cohort. Hepatology (Baltimore, Md.) 2020:hep.31404-31423.

24. Wang Y, Liu S, Liu H, Li W, Lin F, Jiang L, et al. SARS-CoV-2 infection of the liver directly contributes to hepatic impairment in patients with COVID-19. J Hepatol 2020.

25. Bangash MN, Patel JM, Parekh D, Murphy N, Brown RM, Elsharkawy AM, et al. SARS-CoV-2: is the liver merely a bystander to severe disease? Journal of hepatology 2020:1–5.

26. Philips CA, Ahamed R, Augustine P. SARS-CoV-2 related liver impairment - perception may not be the reality. Journal of hepatology 2020:S0168-8278(0120)30344-30345.

27. Dufour DR, Lott JA, Nolte FS, Gretch DR, Koff RS, Seeff LB. Diagnosis and monitoring of hepatic injury. II. Recommendations for use of laboratory tests in screening, diagnosis, and monitoring. Clin Chem 2000;46:2050–2068.

