## Supplementary File for "Abnormal liver tests in admitted patients with SARS-Cov-2 or other respiratory viruses- prognostic similarities and temporal disparities"

Table S1

Figures S1-S4

**Table S1. Univariate analysis of various parameters with a composite outcome of death or mechanical ventilation during hospitalization.**

|  | Influenza | | | | RSV | | | | SARS-Cov-2 | | | |
| --- | --- | --- | --- | --- | --- | --- | --- | --- | --- | --- | --- | --- |
|  | **OR** | **Lower 95% C.L** | **Upper 95% C.L** | **p value** | **OR** | **Lower 95% C.L** | **Upper 95% C.L** | **p value** | **OR** | **Lower 95% C.L** | **Upper 95% C.L** | **p value** |
| Age | 1.04 | 1.02 | 1.06 | **<.0001** | 1.03 | 1.0 | 1.06 | **0.049** | 0.99 | 0.97 | 1.00 | **0.049** |
| Gender | 1.18 | 0.73 | 1.92 | 0.491 | 1.37 | 0.63 | 2.98 | 0.425 | 1.18 | 0.46 | 3.05 | 0.728 |
| BMI | 0.97 | 0.92 | 1.02 | 0.188 | 1.03 | 0.96 | 1.10 | 0.457 | 0.95 | 0.86 | 1.04 | 0.263 |
| CV disease | 1.81 | 1.12 | 2.94 | **0.016** | 1.62 | 0.72 | 3.63 | 0.241 | 2.16 | 0.84 | 5.57 | 0.112 |
| Liver Disease | 1.31 | 0.45 | 3.79 | 0.620 | 0.86 | 0.19 | 3.97 | 0.846 | 0.00 | 0.00 | 0.00 | 0.999 |
| GGT ≥ 40 | 4.76 | 2.74 | 8.28 | **<.0001** | 5.28 | 1.93 | 14.42 | **0.001** | 7.37 | 2.08 | 26.13 | **0.002** |
| GOT ≥ 40 | 5.87 | 3.44 | 10.02 | **<.0001** | 5.70 | 2.48 | 13.12 | **<.0001** | 8.21 | 2.81 | 24.00 | **<.0001** |
| GPT ≥ 40 | 3.66 | 2.23 | 6.01 | **<.0001** | 6.29 | 2.75 | 14.41 | **<.0001** | 7.06 | 2.54 | 19.59 | **<.0001** |
| Bilirubin _max_ | 1.33 | 1.11 | 1.59 | **0.002** | 1.81 | 1.08 | 3.04 | **0.025** | 3.39 | 1.43 | 8.03 | **0.006** |
| Albumin _min_ | 0.20 | 0.13 | 0.28 | **<.0001** | 0.25 | 0.14 | 0.44 | **<.0001** | 0.08 | 0.03 | 0.22 | **<.0001** |
| CRP _max_ | 1.08 | 1.06 | 1.10 | **<.0001** | 1.01 | 1.04 | 1.12 | **<.0001** | 1.12 | 1.07 | 1.17 | **<.0001** |
| Lymph _min_ | 0.12 | 0.05 | 0.29 | **<.0001** | 0.10 | 0.03 | 0.37 | **0.0005** | 1.06 | 0.97 | 1.15 | 0.172 |
| PT/INR _max_ | 1.44 | 1.07 | 1.93 | **0.0168** | 1.41 | 1.02 | 1.94 | **0.0347** | 15.24 | 3.25 | 71.36 | **0.001** |
| Systolic BP _min_ | 0.94 | 0.93 | 0.96 | **<.0001** | 0.95 | 0.93 | 0.97 | **<.0001** | 0.94 | 0.27 | 0.91 | **<.0001** |
| Blood oxygen Saturation _min_ | 0.88 | 0.85 | 0.91 | **<.0001** | 0.88 | 0.82 | 0.95 | **0.0006** | 0.89 | 0.84 | 0.94 | **<.0001** |
| Anti-viral treatment | 1.22 | 0.62 | 2.37 | 0.566 | 1.52 | 0.69 | 3.33 | 0.299 | 1.71 | 0.67 | 4.39 | 0.261 |
| Antibiotics | 6.02 | 2.57 | 14.06 | **<.0001** | 10.35 | 1.37 | 78.03 | **0.0232** | 12.87 | 1.68 | 98.48 | **0.014** |
| Length of stay (days) | 1.06 | 1.04 | 1.08 | **<.0001** | 1.04 | 1.02 | 1.07 | **0.0017** | 1.14 | 1.79 | 1.22 | **<.0001** |
| In Hospital pneumonia | 3.55 | 2.13 | 5.94 | **<.0001** | 6.70 | 2.92 | 15.34 | **<.0001** | 0.00 | 0.00 | 0.00 | 0.999 |
| ICU | 7.82 | 3.13 | 19.51 | **<.0001** | 7.27 | 1.71 | 30.91 | **0.0072** | 22.57 | 5.09 | 100.13 | **<.0001** |

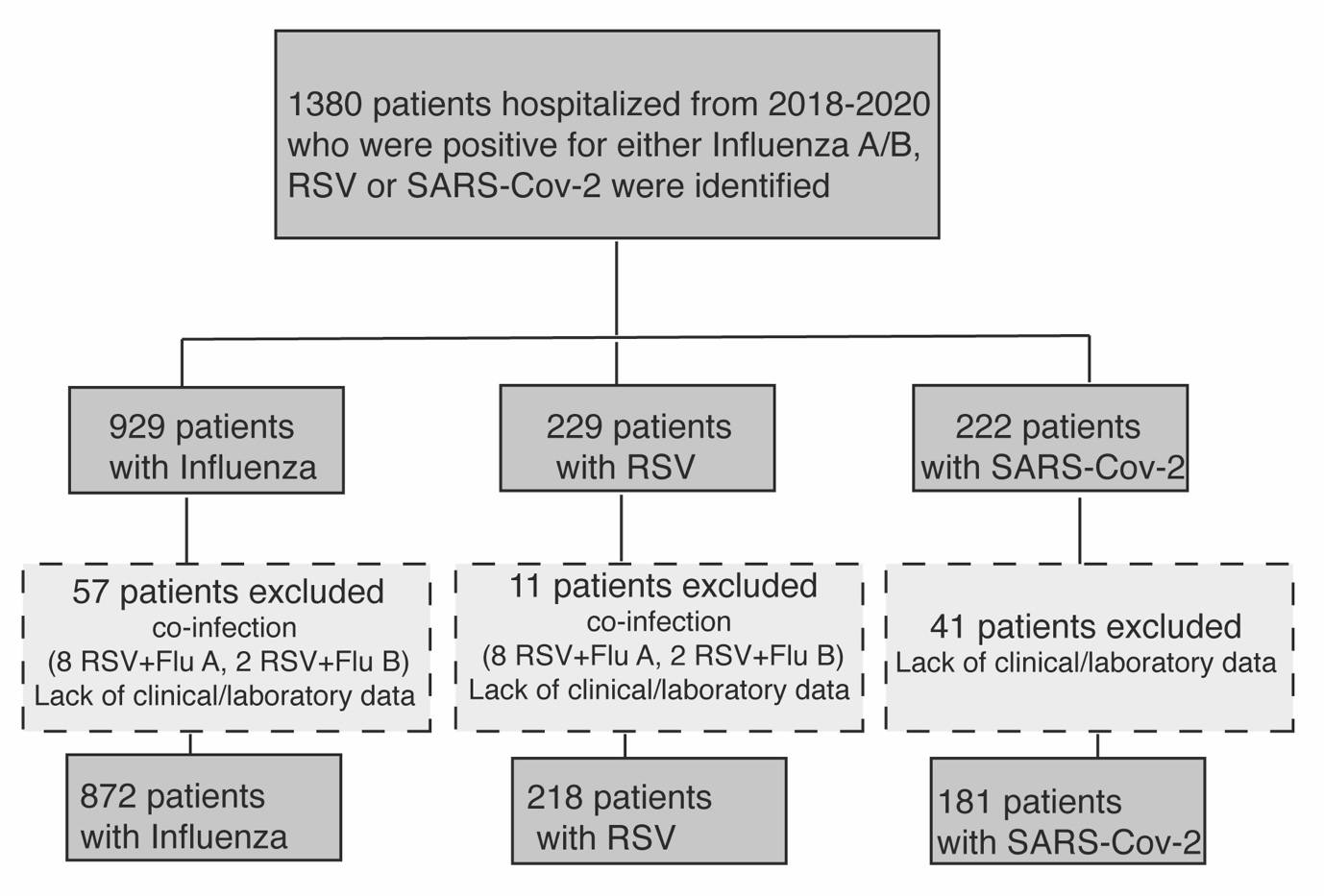

**Fig S1.** Study flow-chart

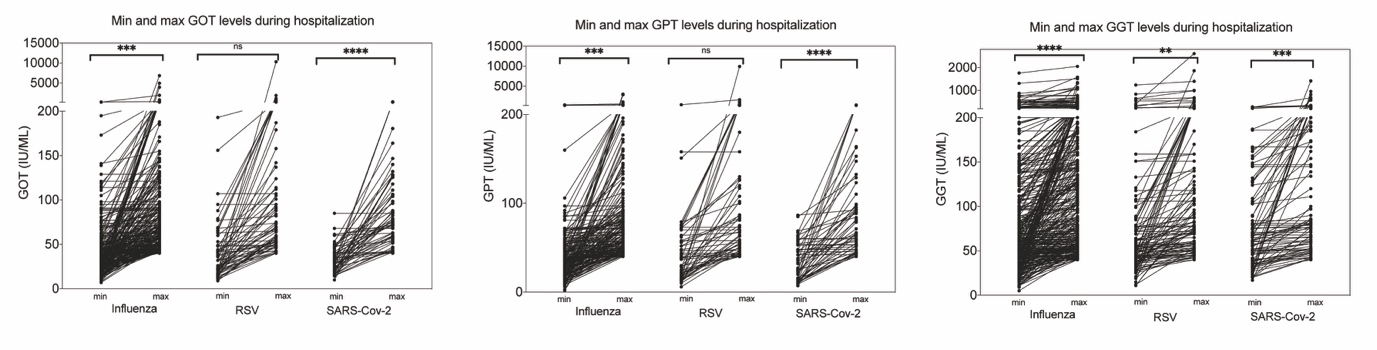

**Fig S2.** Comparison of minimal and maximal levels of liver enzymes during hospitalization among patients with abnormal liver tests (³40) according to the type of infection. Statistical differences were calculated using unpaired t test (ns-non significant, asterisk indicate statistical significance).

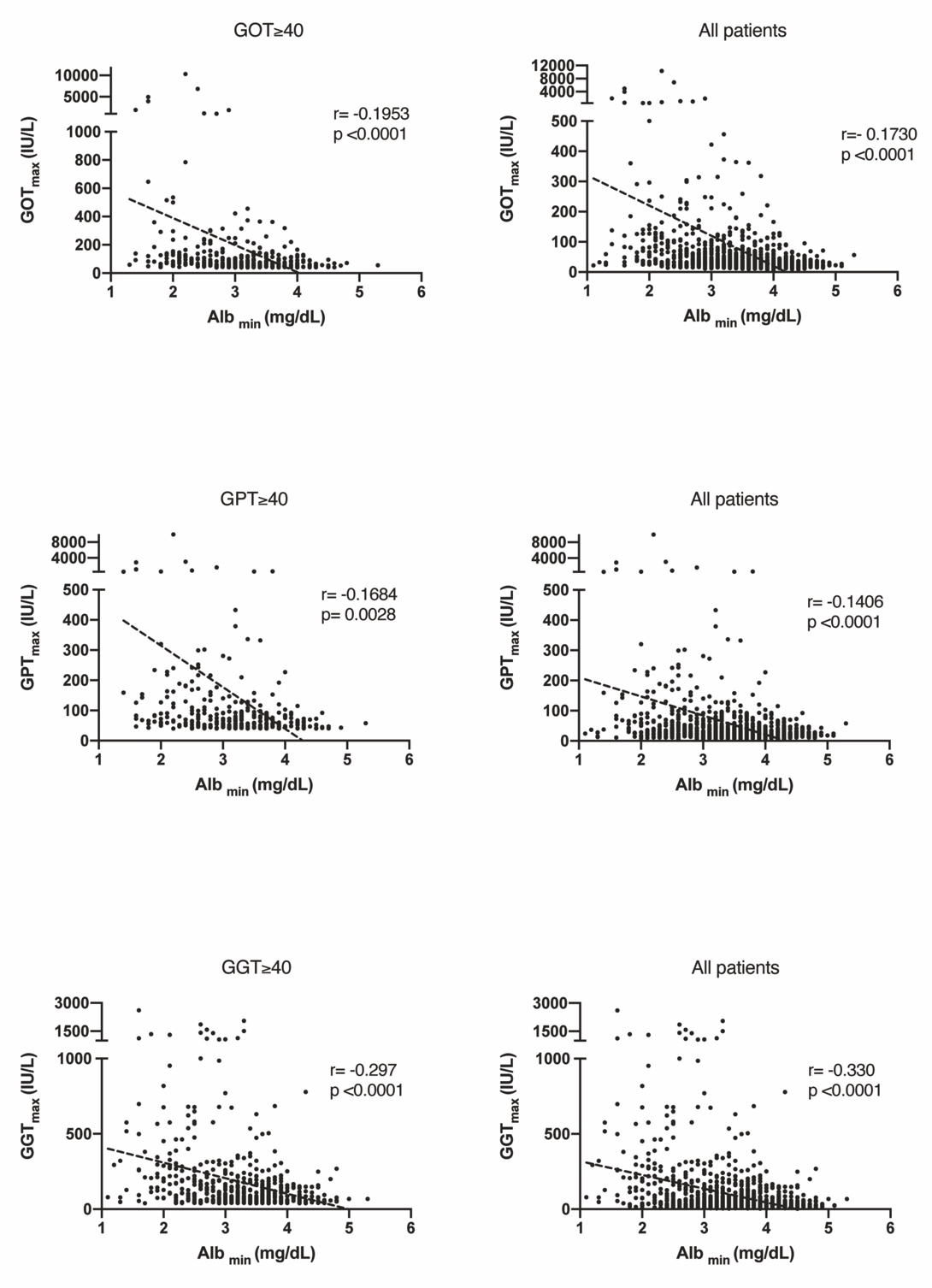

**Fig S3. Correlation curves between serum albumin_min_ and peak LFTs levels during hospitalization.** A pearson’s correlation analysis of the maximal level of each liver enzyme (GPT, GOT or GGT) and minimal albumin level during hospitalization are displayed with the corresponding pearson’s correlation coefficient ( r ) values and their p values. Left panels display the results only for patients with eLFTs (³40) and right panels display the results for all hospitalized patients (regardless of the type of infection).

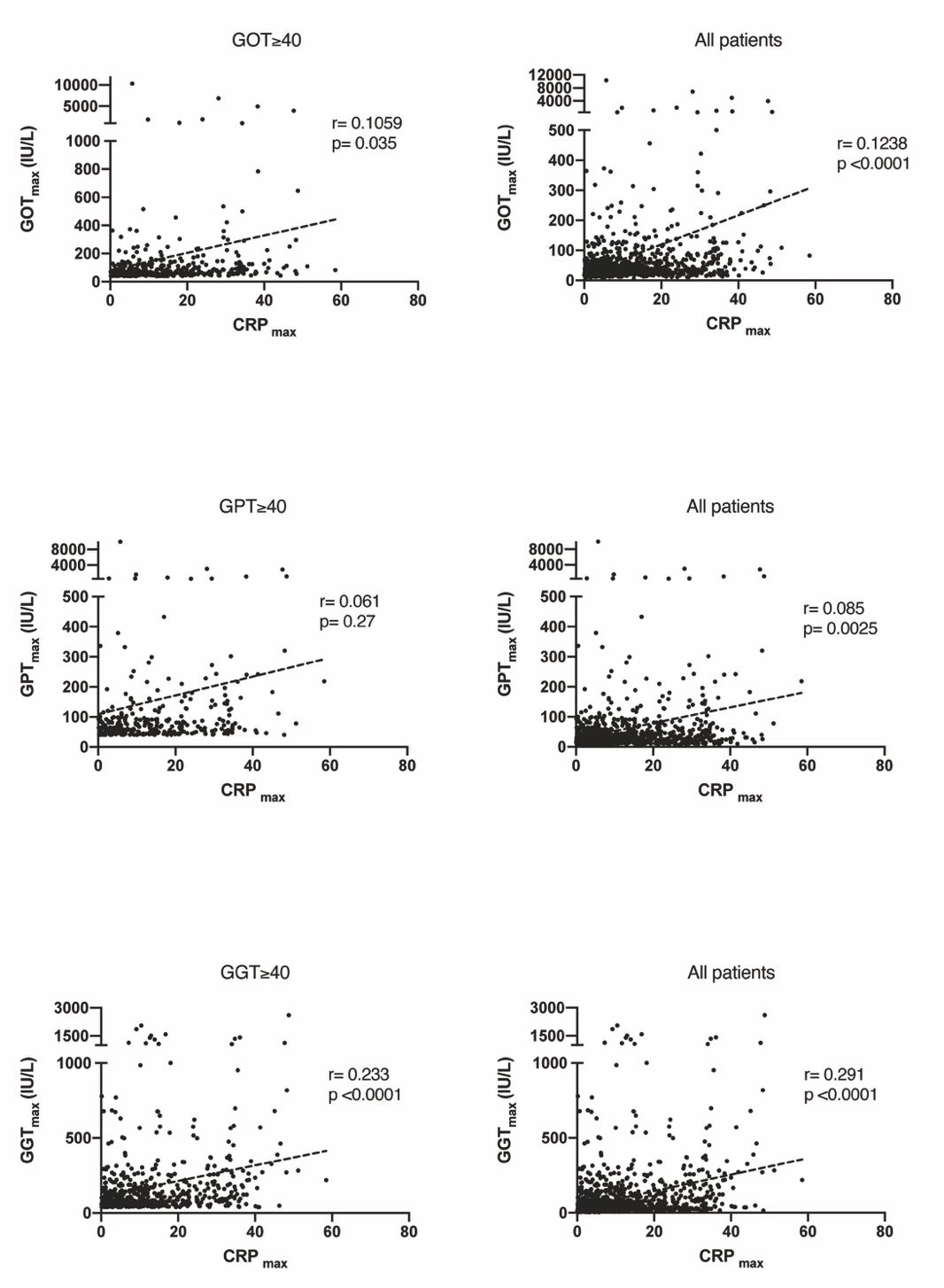

**Fig S4. Correlation curves between serum CRP_max_ and peak LFTs levels during hospitalization.** A pearson’s correlation analysis of the maximal level of each liver enzyme (GPT, GOT or GGT) and maximal CRP level during hospitalization are displayed with the corresponding pearson’s correlation coefficient ( r ) values and their p values. Left panels display the results only for patients with eLFTs (³40) and right panels display the results for all hospitalized patients (regardless of the type of infection).
